# Feasibility Study: Silicone Wristbands for Measuring Pesticides and Flame-Retardants in Farming Older Men

**DOI:** 10.1101/2025.04.17.25326034

**Authors:** Marianne Chanti-Ketterl, Brenda L. Plassman, Christine G. Parks, Heather M. Stapleton

## Abstract

**Background:** Silicone wristbands can detect exposure to multiple chemicals. In the last decade, health studies have used them to assess environmental exposures, but few have included aging men, particularly those actively working with pesticides.

**Objective:** To explore the feasibility, acceptability, and/or receptiveness of older adults to wear a wristband for multiple days while collecting other biomarkers (serum and urine). Additionally, we aimed to assess exposure differences among actively farming pesticide applicators versus non-farming older adults and examine correlations between pesticides and organophosphate ester flame retardants [OPEFR] detected in the wristbands with target serum/urine metabolites.

**Methods:** Fifteen males age 70-plus in North Carolina; 10 farmers, pesticide applicators and 5 non-farmers with no reported pesticide use. Participants wore wristbands for 5 consecutive days 24/hours a day, provided a serum sample on day one and end-of-day urine samples on days 1, 3, and 5. Six pesticides and four OPEFR were measured in wristbands.

**Results:** All participants were receptive to wristbands and wore them for the required time, were compliant in study procedures, and did not report any adverse reactions to the wristbands. In wristbands, atrazine (Geometric means (GM) of specific gravity-corrected levels=14.5 ng/g) and malathion (GM=21.8 ng/g) were detected only in farmers. wristbands GMs were higher in farmers than non-farmers for chlorpyrifos GM difference=408.8 ng/g, and trans-permethrin GM difference=569.2 ng/g. However, wristbands DDT GM was 1.8 ng/g higher in non-farmers than farmers. OPEFRs TCEP and TDCIPP GMs were higher among farmers while TCIPP and TPHP were higher among non-farmers. wristbands pesticide levels and urinary metabolite correlations were weak-to-moderate. A moderate correlation was observed between DDE in wristbands and DDE in serum (r_s_=.44; *p*=0.20).

**Conclusion:** Despite the short test time, wristbands detected multiple chemical exposures regardless of farming status. More research is required to understand the utility of wristbands in assessing pesticide exposures in older populations.

**Impact Statement:** Silicone wristbands are inexpensive, easy to wear passive sampling devices able to capture multiple chemicals providing a snapshot of environmental exposures. Recent research supports wristbands as promising tools to capture flame retardant exposures, but more studies are needed to determine the efficacy of the wristbands for pesticide exposures in older occupational cohorts.

## INTRODUCTION

Silicone wristbands are personal passive sampling devices able to detect multiple environmental bioavailable organic contaminants^1–6^, including specific pesticides^7–12^. Wristbands reflect both dermal and inhaled routes of exposure occurring from both active and passive sources^13,14^ and could be used to monitor complex patterns of exposure in aging populations. Though wristbands have been successfully tested with young and middle-age working adults, mothers and children,^1–12,15–18^, few wristband studies have focused in older adults. Perhaps because with age comes decreases in pharmacokinetic functions which alter the physiological absorption, distribution, metabolism, and elimination of toxic chemicals in the body^19,20^ which may potentially affect the correlation between urine or blood markers and wristbands measures. Nonetheless, characterizing and quantifying chemical exposures in a person’s macro and microenvironments (i.e. farms/homes) regardless of age, may provide important insight into the kind of environmental interventions needed to healthfully age in place.

Some epidemiological studies, such as the Agricultural Health Study (AHS)^21^, aim to characterize the distribution of occupational exposures with health outcomes across the life course. For example, chronic diseases such as hypothyrioidism and depression have been observed at higher rates amongst those most exposed to pesticides in the AHS^19,22,23^. However, non- or inverse associations between the pesticides and other health outcomes have been observed^24^. A limitation of long lasting occupational studies is finding significant associations between the various chemical exposures that occurred decades ago with late-life health outcomes. Thus, testing novel inexpensive chemical exposure assessment methods is needed in these larger epidemiological studies to assess the cohort’s exposures, particularly when addressing aging cohorts with historical lifetime self-report exposure data, but no contemporary exposure measures. This type of data may help unravel whether exposure misclassification exists, or not, in self-report cohorts.

Following prior research and published recommendations ^7–12,15–17,19^ we aimed to test how feasible it would be to use wristbands in older adults participating in the AHS-Memory in Aging Study (AHS-MA) in North Carolina who were going to receive a house visit for biomarker collection. The goal was to not only see how receptive older men would be to using the wristbands, but we also wanted to measure the correlations of chemicals detected in the wristbands with target serum/urine metabolites and to assess differences among actively farming pesticide applicators in AHS-MA versus non-farming older adults.

## METHODS

### Study design and population

The sample included 15 men over the age of 70 from North Carolina; 10 farmers and 5 non-farmers. Farmers were pesticide-applicators identified through the AHS-MA, a sub-study of the AHS, in North Carolina^21^, they were cognitively normal, without any serious health concerns, were scheduled for a blood draw visit as part of the AHS-MA and planning to apply pesticides on the week of the study visit. The farmers were located throughout North Carolina and no two farmers were located on the same farm. All, but one of the farmers, lived on their farm land, the one lived less than 3 miles from the farm. Non-farmers were recruited from the Alzheimer’s Disease Prevention Registry of the Joseph and Kathleen Bryan Alzheimer’s Disease Research Program at Duke University. These were community members who did not endorse the use pesticides or engage in farming activities. Study visits were conducted during November and December, 2018. All participants in the study gave informed consent and received a $25 gift card for participating in the study. Both the AHS-MA study and this pilot study were approved by the IRBs of the National Institute of Environmental Health Sciences and Duke University Medical Center.

### Wristbands, Study Instructions and Exposure Logs

Wristbands were obtained, clean and prepared as previously described^5,25^. Three field blank not deployed, meaning they were not allocated to anyone, were not hand manipulated but opened and left exposed to sampling site environment for less than 5 minutes, and then were wrapped again and brought back to lab. These field blanks were stored at room temperature until extraction for quality assurance and were transported with other deployed wristbands. On day 6, wristbands were collected by research staff and wrapped in aluminum foil, placed in a plastic zip bag and transported back to the lab in a cool cooler.

Participants were instructed to wear the wristband continuously for 24 hours a day for 5 consecutive days. That meant that wristbands were worn all day, including work activities, bathing, and sleeping. Participants were also instructed to collect the day’s last urine on days 1, 3, and 5 during that 5-day study period. The day’s last urine was collected to better capture the day’s exposure to chemicals. Urine was collected in sterile polypropylene specimen containers and immediately stored in a freezer after collection. On day 6, specimens were picked-up and transported to the lab in a cooler with ice. At the lab, for each participant, equal volume aliquots of the three individual urine samples were combined to form one pooled sample before freezing at −20°C for storage.

Last, farmers were asked to complete a daily log including the following questions: (a) Did you have any exposure to pesticides today? (b) Did you mix or touch any pesticides today? (c) Were you out in the field today? Non-farmers were asked daily if they had any known exposure to pesticides on that day.

### Wristbands Targeted Chemicals and Processing

Given the small scope of this study, efficacy testing focused only on chemicals commonly detected in other studies^5,7,12,18^. These included six pesticides widely used in North Carolina and four organophosphate flame retardants and plasticizers (OPFR). The six targeted pesticides included: organophosphate insecticides, chlorpyrifos and malathion; pyrethroid insecticides, cis-permethrin and trans-permethrin; one organochlorine (OC) insecticide, DDT; and one triazine herbicide, atrazine. OPFR included tris (2-chloroethyl) phosphate (TCEP), tris (chloroisopropyl) phosphate (TCIPP), tris (1,3-dichloroisopropyl) phosphate (TDCIPP) and Triphenyl phosphate (TPHP).

Lab personnel were blinded to whether the sample was from a farmer or non-farmer participant. Wristbands were analyzed via gas chromatography high resolution mass spectrometry; details on the specific methods employed are described elsewhere^5,18^. Recoveries of the internal standards ranged from 77 to 144%. Method detection limits were calculated using three times the standard deviation of the three field blank measurements for each analyte. Because no analytes were detected in any of the field blanks, detection limits were based on an instrument signal to noise ratio of 10.

### Urine Processing and Analyses

A 3mL pooled urine sample was shipped on dry ice to a private commercial certified laboratory^26^ overnight for pesticide analyses. Pesticides measured in urine by gas chromatography included atrazine (Primatol A) and four additional herbicides cyanazine, metribuzin, propazine, and simazine. In house, OPFR metabolites were analyzed following methods previously reported in the literature^27–29^. Creatinine was quantified by colorimetry in mg/L units and organophosphate pesticide metabolites were measured by high performance liquid chromatography/Tandem Mass Spectrometry (LC-MS/MS). Performance characteristics were determined as follows: dialkyl phosphate metabolites (DMP [ng/mL], DMTP[ng/mL], DMDTP[ng/mL], DMAP[nmol/L], DMAP creatinine corrected[nmol/g Creat], DEP[ng/mL], DETP[ng/mL], DEDTP[ng/mL], DEAP[nmol/L], DEAP creatinine corrected [nmol/g Creat], DAP[nmol/L], DAP creatinine corrected[nmol/g Creat]). Although not pesticide specific, the following metabolites were targeted as they were included in the most comprehensive lab panel available at the time: chlorpyrifos (Diethyl-phosphate (DEP) and Diethylthio-phosphate=[DEP+DETP]); malathion: (Dimethyl-phosphate, Dimethylthio-phosphate, Dimethyldithio-phosphate= [DMP+DMTP+DMDTP])^30,31^. Urinary OPFR metabolites quantified included: BCIPP (bis(1,3-dichloro-2-propyl) phosphate); BCIPHIPP (1-hydroxyl-2-propyl bis(1-chloro-2-propyl) phosphate); DPHP; BDCIPP; ipPPP (Isopropyl-phenyl phenyl phosphate); and tbPPP (tert-butyl-phenyl phenyl phosphate). These metabolites were associated with their respective parent OPFR in wristbands as follows: TCIPP with BCIPP and BCIPHIPP; TDICPP with BDCIPP (bis(1,3-dichloro-2-propyl phosphate); and TPHP with DPHP(diphenyl phosphate).

### Serum Collection and Analyses

Serum was collected in a 7.5 mL Becton-Dickenson serum separator tube following standard procedures for venipuncture^32^. Serum samples were collected from the farmers per AHS-MA protocol at the time of the first visit and were transported in a cooler with ice from collection site to the University lab. At lab, they were they were spun, aliquoted, and stored at −20°C. Serum aliquots were later shipped on dry ice to the Environmental & Occupational Health Sciences Institute at Rutgers University for analyses of specific organochlorines including Dichlorodiphenyldichloroethylene (DDE) and Dichlorodiphenyltrichloroethane (DDT). DDT was measured via serum p,p’-DDE levels from 1000 uL of serum volume by gas chromatography. Detailed methodology for p,p’-DDE has been previously published^33^.

### Statistical Analyses

To assess the correlations between wristbands chemicals and urine metabolites, we followed standard analytical procedures in the field. Only chemicals with detectable levels in at least 60% of the sample were included in analyses. If a chemical was included for analysis but some samples had values below the minimum detection level (MDL), a value was calculated by taking the MDL of the specific chemical divided by the square root of 2 and normalized to the masses of sample extracted for wristband. Pesticides and OPFR concentrations in wristbands were reported in ng/g. In urine, sum of the metabolite’s values below MDL (ex. Dimethyl Alkyl Phosphates, Sum of Diethyl Alkyl Phosphates) were assigned the highest MDL value from the corresponding individual chemicals. OPFR and DDE in urine were reported in ng/mL.

Given the small sample size, we ran Spearman’s correlations (r_s_) between compounds in wristbands and urine or serum samples and interpreted correlations as weak when r_s_ equaled 0.20–0.39, moderate if 0.40–0.59, strong if 0.60–0.79, and very strong if ≥0.80. Interpretation of correlations should be done with the caveat that these metabolites are not chemical-specific. A sensitivity analysis was done using data from the farmers only to determine if correlations differ when restricting the sample to those who reported the use of pesticides. To determine whether the chemical levels in farmers differed from those in non-farmers, we used Wilcoxon two-sample test exact probability without correction^34^ using NPAR1WAY procedure in SAS. Due to the exploratory nature of this study and the small sample size, p-levels are provided but do not represent statistically powered comparisons. All statistical analyses were performed using SAS software, version 9.4 (SAS Institute, Inc., Cary, NC).

## RESULTS

### Study Characteristics

Most (90%) of farmers were White Non-Hispanic who lived in rural areas of North Carolina. At the time of the study, most farmers tended to cattle and were harvesting or had completed harvesting and were applying pesticides to prepare the soil for next season. Crops harvested included: corn, blueberries, okra, tomatoes, sweet potatoes, pumpkins, and hay. Due to recent hurricane flooding and delays in draining 20% of farmers were not able to apply pesticides as expected. However, they did wear the wristband and visited the fields daily for inspection. It is important to note most of the farmers did not report the specific pesticides they were using, but some of the most commonly reported were: glyphosate and paraquat. Other chemicals reported were: atrazine, permethrin, malathion, 2,4-D, dicamba, 1,3-dichloropropene and chloropicrin (InLine), 2,4-D and triclopyr (Crossbow), and metribuzin. Non-farmers were all White Non-Hispanic, lived in urban areas and reported not performing any type of farm work and not performing activities that required the use of pesticides. Most (80%) were retired and all denied using any kind of pesticides for home use. There were no differences between farmers and non-farmers by age (range 70-90) or creatinine levels (range 674-2158 mg/dL). Sample characteristics and pesticide levels are presented in Table 1.

### Study Acceptability and Practicality

All 15 participants were receptive to using the wristbands as instructed and all agreed to store the urine samples in their freezers until collection time. No difficulties were encountered with phlebotomy. All participants reported wearing the wristbands for the 5 consecutive days and none reported any adverse event and/or inconvenience with it. We did experience one protocol deviation from a non-farmer participant who accidentally discarded the first urine collected when he cleaned the freezer the next day. Thus, the laboratory had to pool the two remaining urine samples for analysis.

### Chemical distributions

#### Wristbands

To calculate a chemical total, each chemical’s molar mass was applied. Farmers’ wristbands presented higher total chemical amounts than non-farmers (Figure 1). Distribution of chemical levels in wristbands are presented in Figure 2-A and 2-B. Specific MDLs, detection frequency (DF) and geometric-means (GM) are shown in Table 1. Overall, among farmers, the medians were substantially lower than GM values for all pesticides, suggesting only one or a few individual farmers were more highly exposed. Of the 10 chemicals measured in wristbands, atrazine and malathion were detected only in farmers. The other 8 chemicals had ≥ 80% detection rates in the full sample. Chlorpyrifos, cis-permethrin and trans-permethrin were detected in everyone (Figure 2-A). The median levels of cis-permethrin and trans-permethrin were lower among farmers than non-farmers; which is notable given the small sample size. The DF for DDE in wristbands was 90% for farmers and 60% for non-farmers. Although the highest DDE level was in one of the non-farmers, the median for DDE was almost 5 times higher in the farmers.

**Figure 1.**
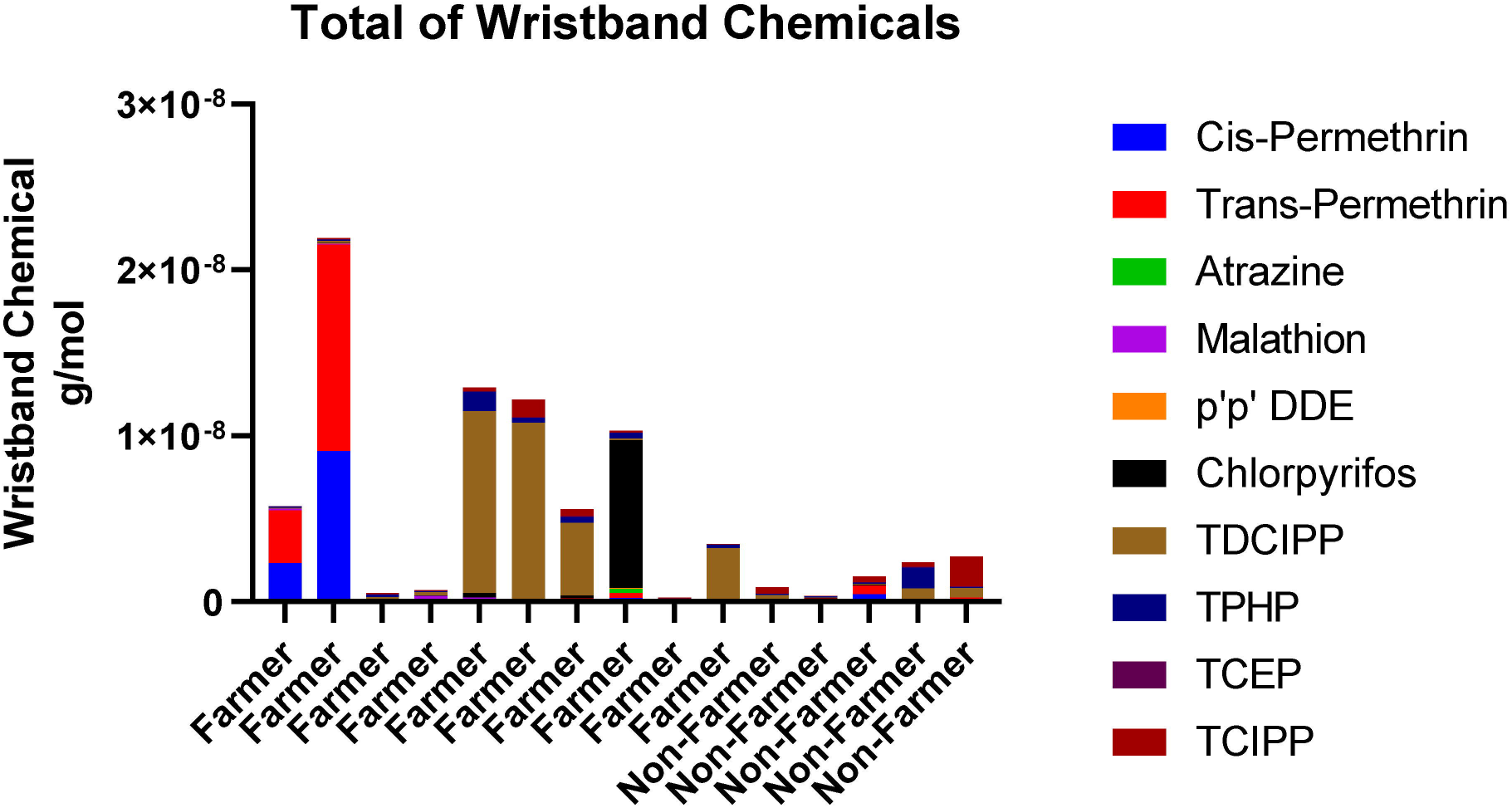
Total of all Chemicals Captured in wristbands for farmers and non-farmers.

**Figure 2.**
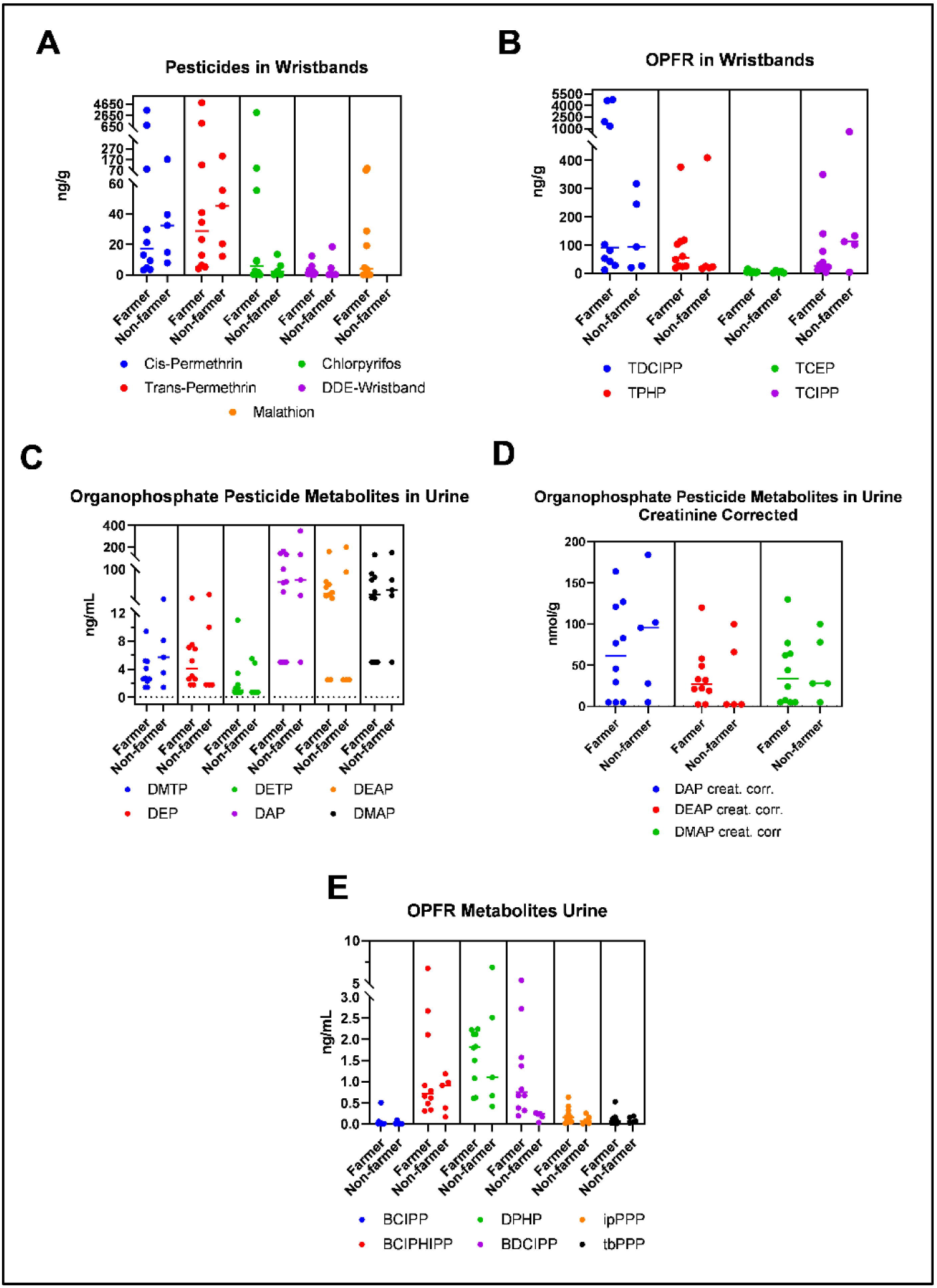
Distribution of chemicals in wristbands and biomarkers in urine for farmers and non-farmers. Note. A. Malathion was detectable >60% only among farmers. B. OPFR (organophosphate flame retardants) detected in wristbands. C. Organophosphate metabolites of pesticides detected in pooled urine samples. D. Organophosphate metabolites of pesticides detected in creatinine corrected urine samples. E. OPFR (organophosphate flame retardants) detected in pooled urine samples.

For the OPFR and plasticizers (Figure 2-B), we obtained 100% DF for TDCIPP and TPHP. TCEP was detected in 100% of farmers and 60% of non-farmers. TCEP and TPHP’s median levels were higher among farmers and TCIPP and TDCIPP were higher among non-farmers; however, the GM for TDCIPP was higher among farmers.

#### Urine

None of the herbicides reached the MDL in any of the urine samples tested. The MDL for atrazine was 20ng/mL and only one sample had a trace level with an estimated concentration ∼15ng/mL. Chemical MDLs are reported in Table 1. Metabolite levels are presented in Figure 2-C, D and E. Only DETP and BCIPP were detected in less than 60% of the samples. Six organophosphate pesticide metabolites were analyzed (Figure 2-C), five were detected in farmers and three in non-farmers. DMTP was the most frequently detected (80% DF) and DETP was the least frequent (40% DF); therefore, DETP was not further analyzed. Although DEP and the sum of DEAP had <60% DF among non-farmers, overall detection among all 15 participants was 60%. Among the farmers, the most frequent organophosphate metabolites were DMTP, DEP and Sum of DEAP with 80% DF. DMTP, Sum of DAP, and Sum of DMAP had 60% or greater DF in both farmers and non-farmers with the GM and maximum level being higher in the non-farmer group than farmers, even after adjusting for creatinine levels (Figure 2-D). However, DEP and the sum of DEAP median levels were higher in farmers than non-farmers.

Five out of the six urinary OPFR metabolites (Figure 2-E) had DF above 60% of the sample. BCIPHIPP, DPHP, and BDCIPP were the most abundant and were detected in the whole sample. With the exception of DPHP, all other OPFRs were higher among farmers. BCIPP was not analyzed because it was only detected in 30% of farmers and 20% of non-farmers. Despite the small sample size, a statistically significant difference was observed between farmers and non-farmers for urinary-BDCIPP where the farmers had almost 8 times higher levels than non-farmers (*p*=.007); but all other urinary levels were not statistically different.

#### Serum DDE

As part of the parent grant scope of work, DDE, the metabolite for DDT was measured only among farmers and was detected in all (Figure 3). Levels were quantified with a median of 0.5 ng/mL and a mean of 1.24 ng/mL [ranged 0.06-5.53 ng/mL].

**Figure 3.**
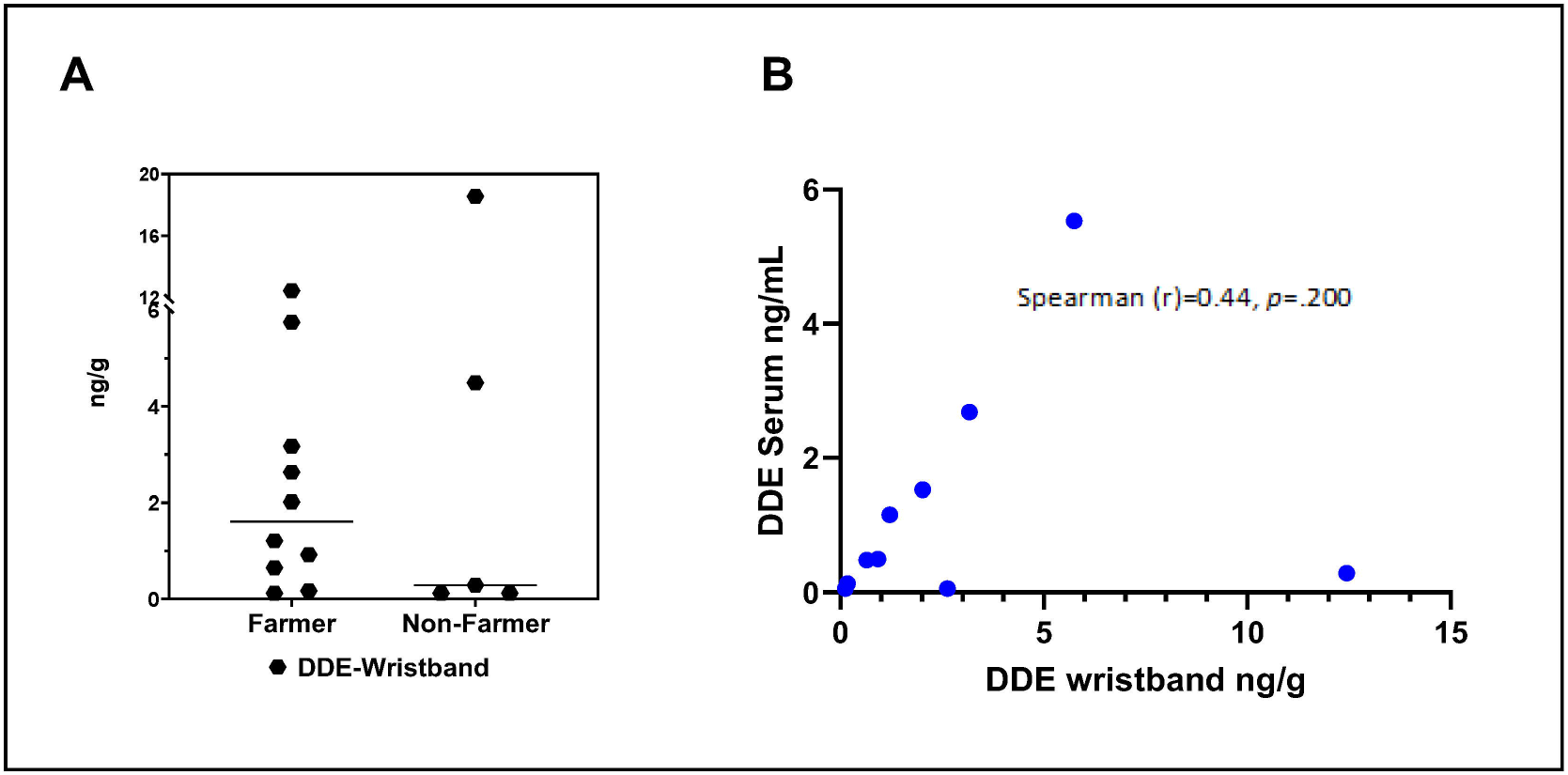
**A.** Distribution of *pp’DDE* in wristbands for farmers and non-farmers. B. Correlation of *pp’DDE* in serum versus *pp’DDE* wristbands in farmers.

### Spearman Correlations

Wristbands with the corresponding urinary metabolite concentrations are presented in Figure 4 and details in supplemental table 1. DETP (urine metabolite for chlorpyrifos), had a DF of 40% in the sample, but we included it in the Spearman’s correlation as it is one of chlorpyrifos major non-specific metabolites. Consequently, wristbands-chlorpyrifos showed a weak correlation with urinary metabolites DETP (r_s_=0.32, *p*=0.248) and a weaker correlation with urinary DEP (r_s_=0.08, *p*=0.776). For the relationship among the OPFR and plasticizers, we observed a weak correlation between wristband-TCIPP and urinary-BCIPHIPP (r_s_=0.35, p=0.203), and moderate correlations for wristband-TDCIPP and wristband-TPHP with their corresponding urinary metabolites, BDCIPP (r_s_=0.49, *p* =0.064) and DPHP (r_s_=0.50, *p*=0.058), respectively. Additional correlations shown on figure 4 may be used to consider relationships between individual chemicals that may occur as mixtures or in close association. For example, among the chemicals detected in wristbands, there was a moderate correlation between chlorpyrifos and TPHP (r_s_=0.48, *p* =0.069).

**Figure 4.**
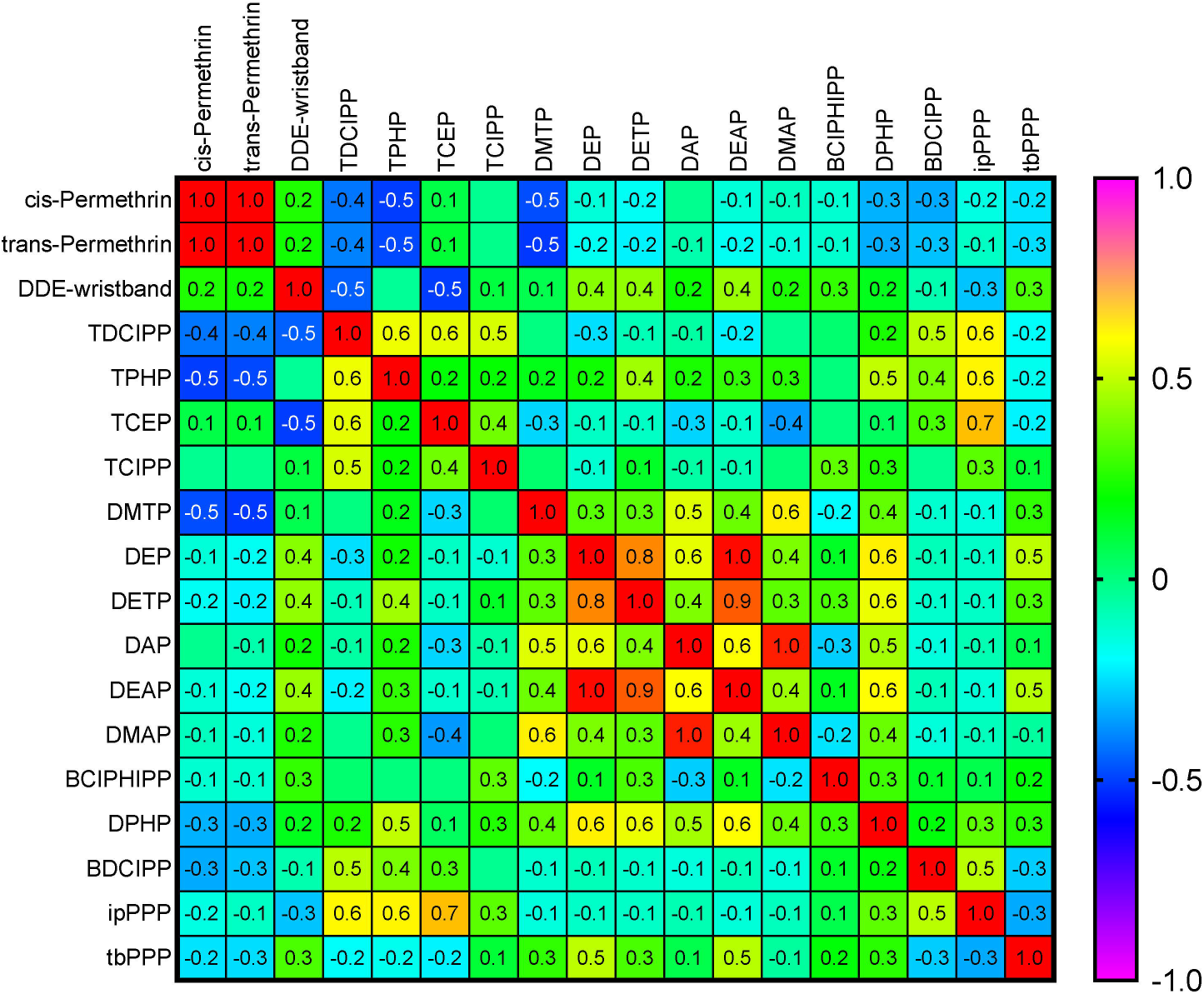
Nonparametric Spearman correlation coefficients for log transformed biomarkers detected in >60% of the sample (n=15).

### Sensitivity analysis among farmers

Given that some of the pesticides were not detected in non-farmers, we ran correlations only among farmers (Supplemental Table 2). Correlations between wristbands-chlorpyrifos and DETP in urine (r_s_=0.36, *p* =0.302) and wristbands-malathion and urinary DMTP (r_s_=0.19, *p* =0.595) were observed. Additionally, a slightly higher correlation between TDCPP and its urinary metabolite BDCIPP (r_s_=0.61, p =0.060) was observed, as well as a moderate correlation between wristbands and serum DDE (*r_s_*=0.44, *p=*.200)(Figure 3-B).

## DISCUSSION

In this study, we aimed to examine if older men would be compliant in using wristbands and to determine if the wristbands would capture chemical exposure differences between actively farming pesticide applicators and non-farming community dwelling older adults. As observed in younger samples^9,10,25^, the use of wristbands in our sample was feasible, well accepted, and the compliance and return rate was 100%. Other studies have reported slightly lower return rates, but those have also had longer deployment periods^1,7,8^.

All six targeted pesticides were detected among farmers, which may suggest that wristbands may work well among older pesticide applicators to objectively capture pesticide exposures, particularly among farmers who appear to be more highly exposed. For example, wristbands captured malathion among 70% of farmers only, which is a much higher detection frequency than previously reported.^9^ However, like others,^7,8,10,11^ we also detected chemicals in the wristbands of non-farmers who reported no exposures. Permethrin, for example, was detected in everyone’s wristbands. Permethrins are pyrethroid pesticides commonly used in many household insecticides, pet products, and are widely detected in fruits and vegetables.^35^ A previous study found higher permethrin levels among participants living within a block of an agricultural field compared to those living further away.^7^ In our study, the mean level of both permethrin pesticides measured were seven to nine times higher among farmers than non-farmers. Chlorpyrifos was also detected among everyone in our sample, but again, at much higher levels than previously reported.^7,9^ Although chlorpyrifos had a voluntary ban from home use products in the United States since 2000,^30^ it remains one of the most globally used and widely studied organophosphates.^30,36–38^

Our study failed to observe significant correlations between organophosphate pesticides and urinary metabolites. This was likely due to a lack specificity. The organophosphate urinary metabolites we measured (i.e. DEP, DETP and DMTP) are not specific to just one pesticide.^38^ For example, we did not see a significant correlation between wristbands chlorpyrifos and DEP or DEPT, but a correlation cannot be ruled out with more specific metabolites. Due to study limitations, we were unable to measure TCP (3,5,6-trichloro-2-pyridinol), which is the specific and primary urinary metabolite for chlorpyrifos^39^. Non-specific metabolites, make it difficult to pin-point the specific parent compounds. For instance, DMTP is a metabolite for 13 different organophosphates^40^, including malathion. In our study, malathion was only capture in farmers’ wristbands, yet DMTP, one of its metabolites, was also detected in 80% of non-farmers urine. Moreover, some non-persistent pesticides with very short half-lives may be difficult to detect, particularly organophosphates, which are mostly excreted in urine within 24 h. Since we collected urine from days 1-3-5 we may have missed their excretion window period. Despite these risk assessment limitations, dialkyl phosphates (DAPs) continue to be used as markers for organophosphate exposures, particularly in clinical and epidemiological studies^41^. Recent studies comparing pre and post DAP levels among individuals who were given an organic diet, showed a significant reduction in DAPs^37,42^. Suggesting that DAPs remain an important organophosphate exposure.

Organochlorines are also of concern as many are highly persistent in soil, sediments, air, and water, as well as in plant and animal tissues. Although banned from the United States since 1972, legacy organochlorine DDT is still in use in many parts of the world to control mosquitos. DDT has a biodegradation half-life in soil between 2-15 years, but the health effects at low environmental levels are unknown.^43^ It is bioaccumulative, an endocrine disruptor, and has been associated with higher risk of cancer, metabolic syndrome, and neurodegenerative and neurobehavioral disorders^44,45^. Commercial mixtures of DDT include *p,p’*-DDT, *o,p’*-DDT, and *p,p’*-DDE as active ingredients. In this study we measured DDE which is the ethylene metabolite of DDT. Our findings parallel previous studies^7,9,11,12^. DDE was detected in 90% of farmer’s and 60% of non-farmer’s wristbands. Overall, wristband DDE’s median level was higher among farmers (1.6 ng/g) and our levels compare to those found in Uruguayan children (1.3 ng/g)^12^. We observed a moderate correlation between DDE in wristbands and DDE in serum (r_s_=.44; *p*=0.20). Despite transnational larger studies using wristbands^46^, we were unable to find another study correlating wristbands and serum DDE levels.

Our study adds to the existing literature^2,4,12,18,46,47^ by reporting levels of OPFR in a much older adult sample. OPFR’s were detected in over 87% of wristbands, with TDCPP and TPHP present in 100%. OPFRs are emerging contaminants widely used in textiles, furniture, and much more as they have been replacing the phased-out polybrominated diphenyl ether flame retardants. We found significant moderate correlations (*rs*=0.50) between TDCIPP and TPHP on wristbands and their urinary metabolites, which parallels previous studies^25,47^. Further, despite our modest sample size, a statistically significant difference in levels of BDCIPP (bis(1,3-dichloro-2-propyl phosphate) and TDCIPP was observed between farmers and non-farmers. Higher wristband levels of TDCIPP have been found in the USA and UK compared to India or China^46^. Our higher wristband and urinary levels of TDCIPP and BDCIPP among farmers may come from environmental exposures which warrants further study.

## Strengths and Limitations

Although our study adds to the current literature by assessing these chemicals in wristbands among active older farming adults and being the first to correlate wristbands DDE levels to serum levels in this population, our study findings should be interpreted with caution. First, our sample size and the number of chemicals analyzed was limited. High resolution mass spectrometry has the capacity to analyze over 1500 chemicals from the wristbands solvent extractions simultaneously; thus, future studies should attempt to analyze larger numbers of chemicals. In addition, due to the small-scale scope of this project, we only included non-specific urinary metabolites. We collected the day’s last urine sample while others have collected the first morning void; this may have contributed to variability across studies due to differences in metabolism rates and short half-lives of some chemicals. It is also worth mentioning, that participants froze the urine samples after each collection, thus urine underwent a freeze/thaw cycle prior to analysis. Last, our results likely do not represent peak exposures during a typical year since they were collected towards the end of the harvest season in fall and under challenging meteorological conditions. During the data collection part of this study, North Carolina experienced a hurricane followed shortly by a tropical storm causing flooded lands and crop loss which lasted well into the end of the year. Because chemicals in the environment and in urine may vary by season or work performed, future studies should use multiple wristbands measures over an extended period of time to adequately capture a person’s overall exposure.

## Conclusion

More research is required to understand whether and how well wristbands can capture aging farmers’ exposures to pesticides in pesticide application and non-application scenarios, and whether they can inform exposure-outcome assessments in epidemiology. Larger studies with comprehensive pesticide absorption/kinetic information, and assessments for more pesticides and specific urine metabolite combinations are needed to determine whether they can objectively differentiate exposures among active farmers. While wristbands offer a more convenient and less expensive tool to measure exposures longitudinally within an individual as compared to collecting urine or blood, much more work is needed to demonstrate usefulness for pesticide exposure classification in active farming population epidemiology studies.

## Data Availability

All data produced in the present study are available upon reasonable request to the authors.

## CRediT author statement

**Chanti-Ketterl:** Conceptualization, Methodology, Formal analysis, Investigation, Writing- Original draft preparation, Visualization, Funding acquisition. **Plassman**: Conceptualization, Funding acquisition, Project administration, Resources, Writing- Reviewing and Editing. **Parks**: Writing- Reviewing and Editing. **Stapleton**: Conceptualization, Methodology, Formal analysis, Funding acquisition, Writing- Reviewing and Editing.

## Acknowledgements

The authors acknowledge Julie Fleenor for recruitment and coordination of the study. The authors also thank Nicholas Herkert and Sharon Zang for processing and analyzing the wristbands samples.

